# High SARS-CoV-2 seroprevalence in Lagos, Nigeria with robust antibody and cellular responses

**DOI:** 10.1101/2022.11.30.22282833

**Authors:** Sulaimon Akanmu, Bobby Brooke Herrera, Beth Chaplin, Sade Ogunsola, Akin Osibogun, Fatima Onawoga, Sarah John-Olabode, Iorhen E. Akase, Augustina Nwosu, Donald J Hamel, Charlotte A Chang, Phyllis J Kanki

## Abstract

**Background:** Early evidence suggested that the impact of the COVID-19 pandemic was less severe in Africa compared to other parts of the world. However, more recent studies indicate higher SARS-CoV-2 infection and COVID-19 mortality rates on the continent than previously documented. Research is needed to better understand SARS-CoV-2 seroprevalence and immunity in Africa.

**Methods:** Our collaboration with the Lagos State COVID-19 Taskforce, enabled secondary analyses of immune responses in healthcare workers (HCWs) and Oxford/AstraZeneca COVID-19 vaccine recipients from the general population across 5 local government areas (LGAs) in Lagos State, Nigeria. Western blots were used to simultaneously detect SARS-CoV-2 spike and nucleocapsid (N) antibodies and stimulation of peripheral blood mononuclear cells with N followed by an IFN-γ ELISA was used to examine T cell responses.

**Findings:** Antibody data demonstrated high SARS-CoV-2 seroprevalence of 71.6% (96/134) in HCWs and 54.8% (63/115) in the general population. Antibodies directed to only SARS-CoV-2 N, suggesting pre-existing coronavirus immunity, were seen in 10.4% (14/134) of HCWs and 20.0% (23/115) of the general population. T cell data showed that IFN-γ responses against SARS-CoV-2 N were robust in detecting exposure to the virus, demonstrating 87.5% sensitivity and 92.3% specificity.

**Interpretation:** These results have important implications for understanding the paradoxical high SARS-CoV-2 infection with low mortality rate in Africa as compared to other parts of the world, as well as for the development of T cell-based diagnostics and vaccines.

**Funding:** Harvard University, Motsepe Presidential Research Accelerator Fund for Africa

## Introduction

Early projections of SARS-CoV-2 spread in Africa fueled fears that the healthcare infrastructure on the continent would be ill prepared to cope with the anticipated COVID-19 hospitalizations and deaths (1). However, as of October 2022, 12.1 million COVID-19 cases and 256,000 deaths have been reported in Africa, representing ∼2% and ∼4% of global statistics, respectively. Early studies indicated a less severe epidemiological picture of COVID-19 in Africa, with a higher proportion of cases resulting in asymptomatic infections and lower mortality compared to other parts of the world (2). Several factors may help explain these phenomena including the younger age demographics with stronger immune status, climate and environmental factors, and weak health systems for reporting (3-5).

In Africa, SARS-CoV-2 prevalence studies conducted in blood donors, healthcare workers (HCWs), pregnant women, and others have described varying rates during different time periods, ranging from 0.4% in Cape Verde (June-July 2020) to more than 49% in Kenya (August 2020-October 2021) and 79% across 12 states in Nigeria (June-August 2021) (6-8). In most cases, the reported prevalence of antibodies against SARS-CoV-2 was several orders of magnitude higher than would be expected from PCR confirmed cases. However, more recent evidence indicates an underestimation of COVID-19’s impact in Africa. In Zambia, postmortem surveillance conducted between June 15 and October 1, 2020, detected SARS-CoV-2 RNA in ∼20% of subjects sampled within 48 hours of death, and only 2 of the 70 had been diagnosed with SARS-CoV-2 prior to death (9). A follow-up study showed that during peak transmission periods, approximately 90% of all deceased individuals tested positive for COVID-19 (10). While these studies demonstrate under-reporting of SARS-CoV-2 infection, there remains a lack of evidence for major increases in mortality, at least suggestive of lower pathogenic impact.

The humoral response to SARS-CoV-2 has been the primary focus of most studies conducted in Africa. A study of pre-pandemic samples in Gabon and Senegal, demonstrated significant pre-existing immunity based on antibodies directed to the SARS-CoV-2 spike (S) and nucleocapsid (N) proteins, in contrast to samples from Canada, Brazil and Denmark (11). However, Gabonese sera with or without antibodies to SARS-CoV-2 N were unable to neutralize the virus in vitro or in mouse infection studies. These data, along with other published studies confirm pre-existing humoral responses to SARS-CoV-2 that may be qualitatively and quantitatively distinct in Africa.

The study of T cell responses has also been used in research and clinical settings during the COVID-19 pandemic to provide further insights into the immune response to infection and/or vaccination. In contrast to humoral responses, T cell responses to human coronaviruses, including SARS-CoV-1 and SARS-CoV-2, may be long-lasting even many years after infection (12, 13). SARS-CoV-2-specific T cells were maintained at 6-9 months following primary infection, indicating that T cell immunity may persist beyond antibody responses (12). Preclinical development of SARS-CoV-2 vaccines have demonstrated T cell responses accompanying antibody responses and clinical studies are now demonstrating similar timelines following vaccination (14). These findings, together with studies that have demonstrated a role for T cells in viral clearance, suggest that cell-mediated immune responses may be an important component of protection against SARS-CoV-2 (15). Here we report the results of antibody and T cell analyses using Nigerian samples from HCWs and a group of vaccine recipients from the general population prospectively followed post-vaccination. To our knowledge, this is the first study to examine both the antibody and T cell response to SARS-CoV-2 in West Africans.

## Methods

### Study Population and Ethics Statement

Our original study in early 2021 proposed to only recruit HCWs, a population with high-risk occupational exposure to COVID-19. However, the roll-out of COVID-19 vaccines in Nigeria at the end of March 2021 enabled study of vaccine recipients from communities also from Lagos state. The Oxford AstraZeneca vaccine, with its low cost and simple refrigeration requirements enabled equitable access for low- and middle-income countries and was the first non-profit vaccine to report efficacy. The recommended initial course is two doses, administered intramuscularly with an interval of 8–12 weeks between doses.

The HCW cohort was recruited from the Lagos University Teaching Hospital (LUTH). Study participation included a brief clinical examination, questionnaire and venous blood sample. HCWs included some with documented prior COVID-19 infection by PCR. HCW samples were collected between March and October 2021.

The Lagos State Vaccine study population included individuals from the general population from Agbowa, Amuwo, Ikorodu, Iwaya, and Oshodi, Lagos State, Nigeria, all of whom, at the time of enrollment, had no documented history of SARS-CoV-2 infection and were administered the first dose of the Oxford/AstraZeneca COVID-19 vaccine between March to June 2021. A brief questionnaire on demographics and exposure was administered at baseline, and venous blood samples were collected at baseline (immediately before vaccination), 7-, 14-, and 84-days after vaccination for immune response testing.

All individuals from each of the 2 cohorts provided written informed consent for the collection of samples and data. The HCW study received ethical clearance from the Harvard T.H. Chan School of Public Health Institutional Review Board (IRB, Protocol #: IRB-21-0329) and the LUTH Health Research Ethics Committee (HREC, Protocol #: ADM/DCST/HREC/APP/4192). The original Lagos State COVID-19 Vaccine study was reviewed and approved by the LUTH HREC (Protocol #: ADM/DCST/HREC/APP/4207), and the secondary analysis of these samples was determined not human subjects research by the Harvard IRB (Protocol #: IRB21-1350).

### Viral Lysate

Briefly, Vero cells infected with SARS-CoV-2 (Isolate USA-WA1/2020, BEI Resources NR-52281) were lysed when cytopathic effect were observed in 20% of cells with lysis buffer containing protease inhibitors, followed by centrifugation at 170,000 x g at 4°C for 90 minutes to obtain cell lysates. Aliquots of cell lysates were purified by sucrose gradient.

### Western Blot

Aliquots of cell lysates were added to nonreducing buffer (final concentrations of 2% SDS, 0.5 M Tris pH 6.8, 20% glycerol, 0.001% bromophenol blue) and subjected to 12% PAGE and Western blot analysis using patient serum (1:250) as primary antibody and anti-human IgG horseradish peroxidase (HRP) (1:2,000; ThermoFisher Scientific, Waltham, MA) as secondary antibody. Visualization was performed using Metal Enhanced DAB Substrate Kit (ThermoFisher Scientific, Waltham, MA) per the manufacturer’s instructions.

### Image analysis

Western blots were analyzed using image processing software, ImageJ (NIH), to machine-read and quantify SARS-CoV-2 S antibody signals. The average pixel intensity was quantified at the Western blot SARS-CoV-2 S band, background areas, and a control band. The background-adjusted SARS-CoV-2 S band signal was then normalized to the background-subtracted control band and expressed as % of control.

### T cell stimulation

From each individual, 3 ml whole blood was collected in a vacutainer tube treated with lithium or sodium heparin (BD, Franklin Lakes, NJ), and tubes were inverted 10 times to ensure that the blood mixed thoroughly with the anticoagulant. 1 ml whole blood was then pipetted into three MASI Stimulator Tubes (Mir Biosciences, Boston, MA) either coated with SARS-CoV-2 N, positive control (phytohemagglutinin), or negative control (PBS). All tubes were then inverted 25 times to ensure that the whole blood mixed thoroughly along the inner walls of the MASI Stimulator Tubes. The MASI Stimulator Tubes were incubated in a humidified incubator at 37°C with 5% CO_2_ for 18-22 hours. After the incubation step, the MASI Stimulator Tubes were centrifuged at 2,500 x g for 5 minutes and the supernatant was aliquoted into 1 ml cryotubes and immediately processed by the IFN-γ ELISA.

### IFN-γ ELISA

Supernatants (100 μl) collected from the MASI Stimulator Tubes were screened for the presence of human IFN-γ by the MASI-COVID enzyme-linked immunosorbent assay (ELISA) (Mir Biosciences, Boston, MA), according to the manufacturer’s instructions. The presence of IFN-γ was captured using a microplate reader (optical density 450 nm). Assay performances were monitored using internal controls and cutoffs were determined as specified by the manufacturer for the kit.

### Statistics

For both the HCW and general population vaccine recipient cohorts, we calculated baseline SARS-CoV-2 S and N antibody seroprevalence determined by Western blot as percentage of total. Using these seroprevalence categories, we evaluated T cell responses after stimulation with SARS-CoV-2 N by plotting IFN-γ ELISA signal in a subset of HCW baseline samples and 7-day post-vaccination general population samples. Additionally, for the vaccine recipient cohort, we used the Western blot and image analysis to evaluate boosting of SARS-CoV-2 S antibodies in sequential samples post-vaccination by calculating mean S signal and change over time. The mean S antibody signal between groups was compared by T-test. Finally, we calculated sensitivity and specificity of IFN-γ responses against SARS-CoV-2 N. All statistics and plots were generated using Prism (version 9.0.0).

## Results

Our study included a cohort of 134 HCWs working at LUTH. Forty HCWs previously tested positive for COVID-19 by RT-PCR and 94 did not have history of documented SARS-CoV-2 infection. (Table 1). Our study also included a second cohort of 126 individuals from the general population across 5 local government areas of Lagos State: 25 from Agbowa, 26 from Amuwo, 25 from Ikorodu, 25 from Iwaya, and 25 from Oshodi (Table 1). At the time of enrollment, referred to throughout as baseline, a blood sample was collected from each individual, immediately prior to administration of the first dose of the Oxford/AstraZeneca COVID-19 vaccine. For these individuals, baseline and follow-up samples were collected between March and August 2021.

**Table 1.** SARS-CoV-2 antibody seroprevalence among healthcare workers and the general population.

| Cohort | Number | Antibody Seroprevalence at Baseline |  |  |  |
| --- | --- | --- | --- | --- | --- |
|  |  | SARS-CoV-2: S+N<br>Number Positive (%) | SARS-CoV-2: S-only<br>Number Positive (%) | SARS-CoV-2: N-only<br>Number Positive (%) | Seronegative<br>Number Positive (%) |
| Healthcare Workers |  |  |  |  |  |
| COVID-19 convalescent | 40 | 38 (95.0) | 2 (5.0) | - | - |
| No history of COVID-19 | 94 | 55 (58.5) | 1 (1.1) | 14 (14.9) | 24 (25.5) |
| Overall | 134 | 93 (69.4) | 3 (2.2) | 14 (10.4) | 24 (17.9) |
| General Population –<br>Vaccine Recipients |  |  |  |  |  |
| Agbowa | 24 | 15 (62.5) | 1 (4.2) | 5 (20.8) | 3 (12.5) |
| Amuwo | 23 | 8 (34.8) | 1 (4.4) | 5 (21.7) | 9 (39.1) |
| Ikorodu | 24 | 16 (66.7) | - | 4 (16.7) | 4 (16.7) |
| Iwaya | 22 | 9 (40.9) | 1 (4.6) | 4 (18.2) | 8 (36.4) |
| Oshodi | 22 | 12 (54.6) | - | 5 (22.7) | 5 (22.7) |
| All sites | 115 | 60 (52.2) | 3 (2.6) | 23 (20.0) | 29 (25.2) |

We analyzed the SARS-CoV-2 seroprevalence based on antibodies directed against SARS-CoV-2 S+N or S-only. The overall seroprevalence of SARS-CoV-2 in HCWs was 71.6% (96/134) (Table 1). Of HCWs with previous RT-PCR confirmation of COVID-19, 100% (40/40) had SARS-CoV-2 S+N or S-only antibodies (Table 1). Of HCWs without any prior history, 58.5% (55/94) had SARS-CoV-2 S+N antibodies, whereas 1.1% (1/94) had S-only antibodies (Table 1). We then examined the T cell response against SARS-CoV-2 N. In HCWs with SARS-CoV-2 S+N antibodies, 82.6% (19/23) had a T cell response against N (Fig. 1A).

**Fig. 1.**
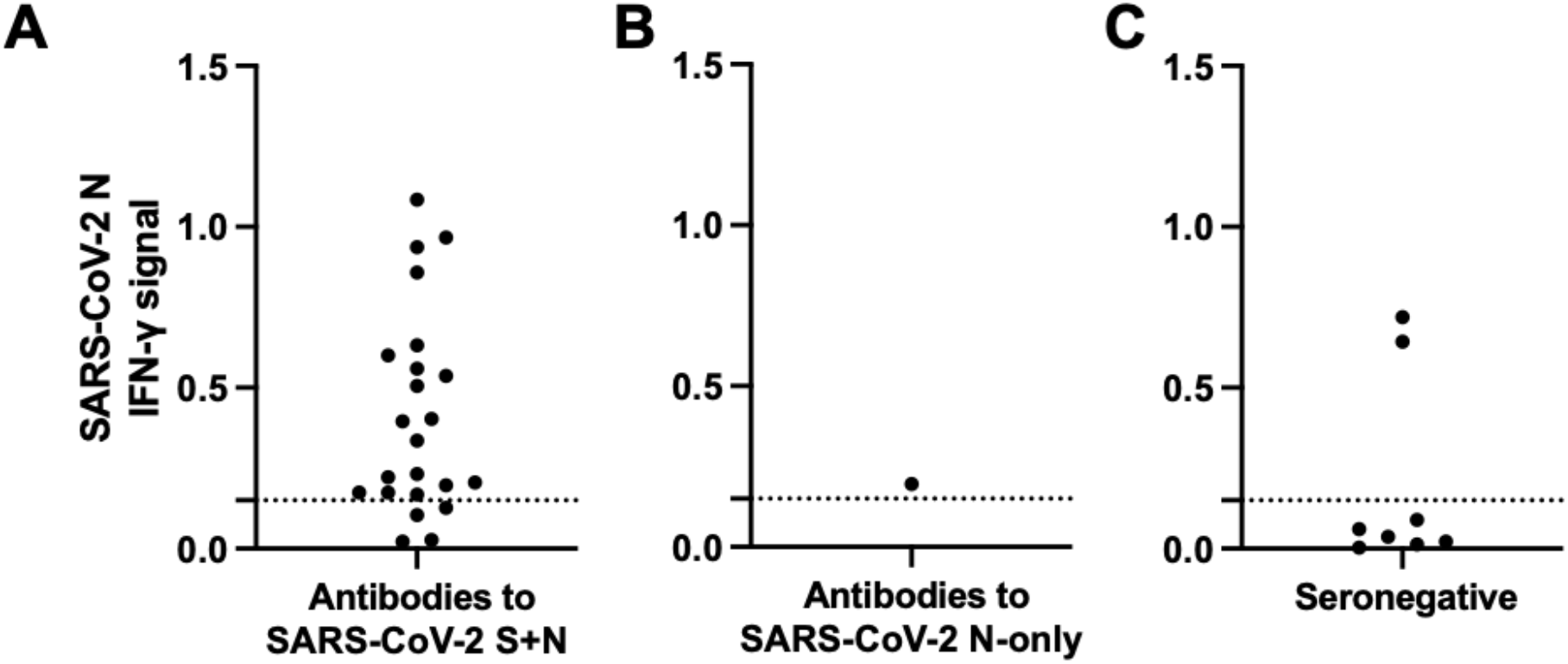
T cell responses to SARS-CoV-2 nucleocapsid (N) in healthcare workers (HCWs). Whole blood samples were stimulated with SARS-CoV-2 N, then supernatants were processed via an IFN-γ enzyme-linked immunosorbent assay for healthcare workers who had antibodies to A) SARS-CoV-2 S+N, B) N-only, or C) were seronegative. Responses are expressed as an arbitrary unit (IFN-γ signal) based on an OD_405_ measurement. Dashed line, assay cutoff.

We next examined HCWs with antibodies directed against SARS-CoV-2 N-only, suggestive of pre-existing coronavirus immunity. There was reactivity to N-only in 10.4% (14/134) of HCWs, all with no history of PCR-confirmed SARS-CoV-2 infection (Table 1). Of those we tested for T cell responses, 100% (1/1) had a T cell response against SARS-CoV-2 N (Fig. 1B). Additionally, 17.9% (24/134) of HCWs were seronegative to SARS-CoV-2. Of those we tested, 75% (6/8) did not have a T cell response against SARS-CoV-2 N (Table 1, Fig. 1C).

We similarly analyzed the SARS-CoV-2 antibody profiles in the general population vaccine recipients. At baseline, the overall SARS-CoV-2 seroprevalence was 54.8% (63/115), with reactivity to SARS-CoV-2 N-only in 20% (23/115), suggestive of pre-existing coronavirus immunity (Table 1). 25.2% of the general population vaccine recipients were seronegative (29/115). Across the five local government areas, SARS-CoV-2 seroprevalence ranged from 34.8% in Amuwo to 66.7% in Ikorodu. The reactivity to SARS CoV-2 N-only was more consistent across the local government areas, ranging from 16.7% in Ikorodu to 22.7% in Oshodi (Table 1).

Given that the Oxford/AstraZeneca COVID-19 vaccine is designed to generate immunity against the SARS-CoV-2 S, we next examined the evolution of SARS-CoV-2 S antibodies post-vaccination. Most individuals, regardless of their SARS-CoV-2 antibody status at baseline, had detectable S antibodies post-vaccination (Fig. 2A-E). However, the potency of the SARS-CoV-2 S antibody response differed between baseline antibody profile groups. The S antibody response was significantly stronger in individuals with baseline SARS-CoV-2 S+N antibodies compared to those with N-only antibodies or seronegatives at 7- (p < 0.0001 or p < 0.0003, respectively) and 14-days (p < 0.002 or p < 0.02, respectively) post-vaccination (Fig. 2F). For the SARS-CoV-2 S+N baseline group, the S antibody response remained significantly stronger only compared to the seronegative group at 84-days post-vaccination (p < 0.03) (Fig. 2F). The SARS-CoV-2 S-only baseline group had a significantly stronger S antibody response compared to the N-only baseline group only at 7-days post-vaccination (p < 0.005) (Fig. 2F). The SARS-CoV-2 N-only baseline group had a significantly stronger S antibody response compared to the seronegative baseline group only at 84-days post-vaccination (p < 0.004) (Fig. 2F). Also, 6 of 115 individuals (5.2%), all of whom were seronegative at baseline, failed to develop SARS-CoV-2 S antibodies 4 weeks after the second vaccine dose (Fig. 2D-E).

**Fig. 2.**
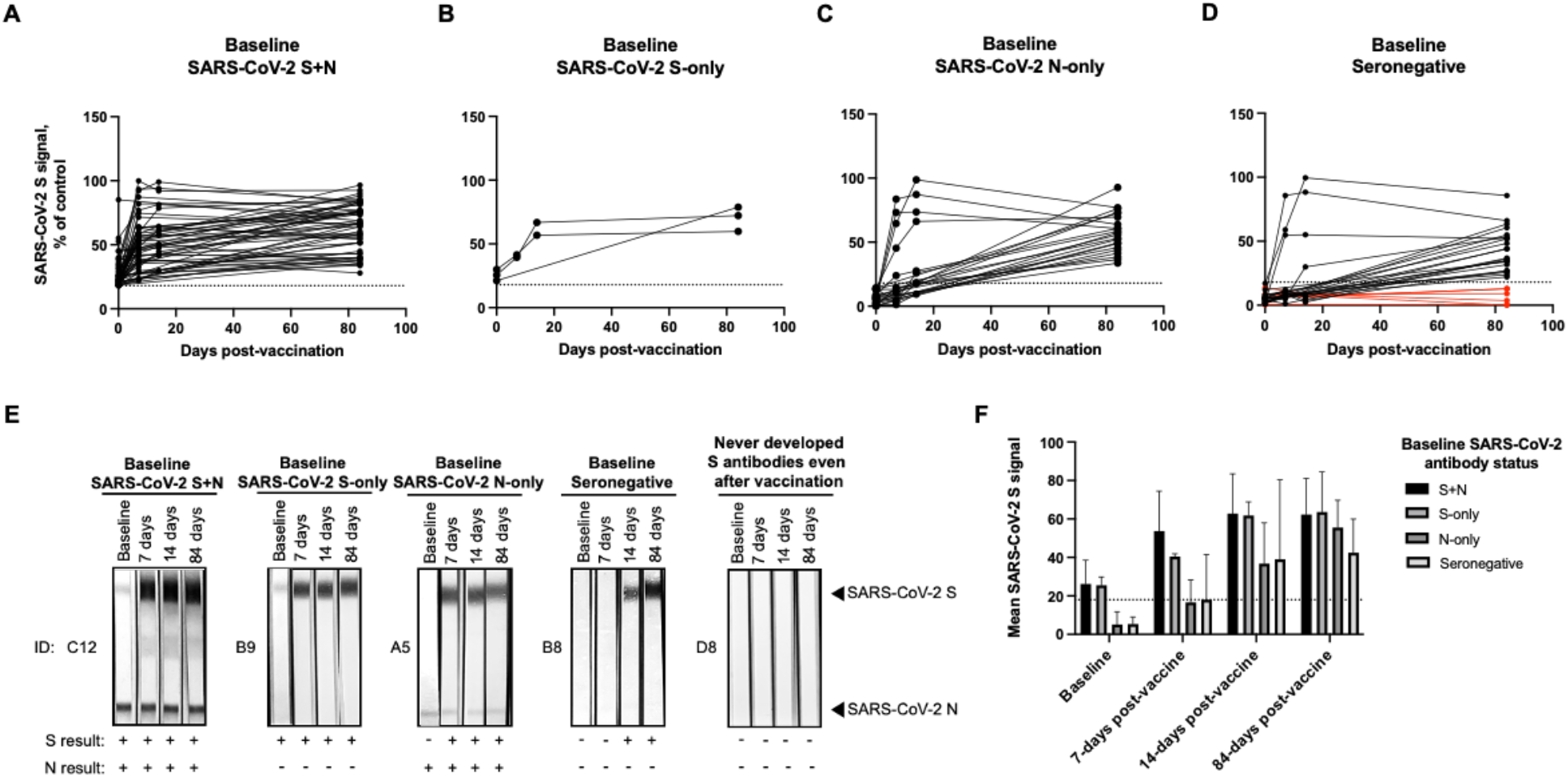
Evolution and boosting of SARS-CoV-2 spike (S) in individuals in the general population post-vaccination. Sera sequentially collected from vaccine recipients in the general population were subjected to Western blot analysis. The post-vaccination evolution of SARS-CoV-2 S antibodies for each individual with baseline antibodies to A) SARS-CoV-2 S+N, B) S-only, C) N-only, or D) who were seronegative (red lines represent individuals who never developed S antibodies even after two vaccine rounds) determined by image analysis where the S antibody signal was calculated and plotted as a % of control. The x-axis corresponds to number of days post-vaccination. The y-axis corresponds to background subtracted SARS-CoV-2 S signal normalized to the control line for each Western blot. Dashed line, cutoff. E) The evolution of SARS-CoV-2 S antibodies was determined by image analysis where the S antibody signals were calculated and plotted as the mean S signal by time period for each baseline antibody profile group. Dashed line, cutoff. F) Representative image of Western blots for groups of individuals.

Since some individuals already had SARS-CoV-2 S antibodies at baseline, suggesting prior SARS CoV-2 infection, we also examined the levels of S boosting post-vaccination. In individuals with SARS-CoV-2 S+N antibodies at baseline, 74.4% of S boosting occurred within 7-days post vaccination, followed by 24.5% and 1.2% within 14- and 84-days post-vaccination, respectively (Fig. 3). For individuals with S-only antibodies or were seronegative at baseline, ∼36.6% of S boosting occurred within 7-days post-vaccination, followed by 56.4% and ∼7% within 14- and 84-days post-vaccination, respectively (Fig. 3). S boosting in individuals with SARS-CoV-2 N-only antibodies at baseline differed compared to other groups, with relatively consistent levels between time periods. For these individuals, 22.9%, 39.9%, and 37.2% of S boosting occurred within 7-, 14-, and 84-days post-vaccination, respectively (Fig. 3).

**Fig. 3.**
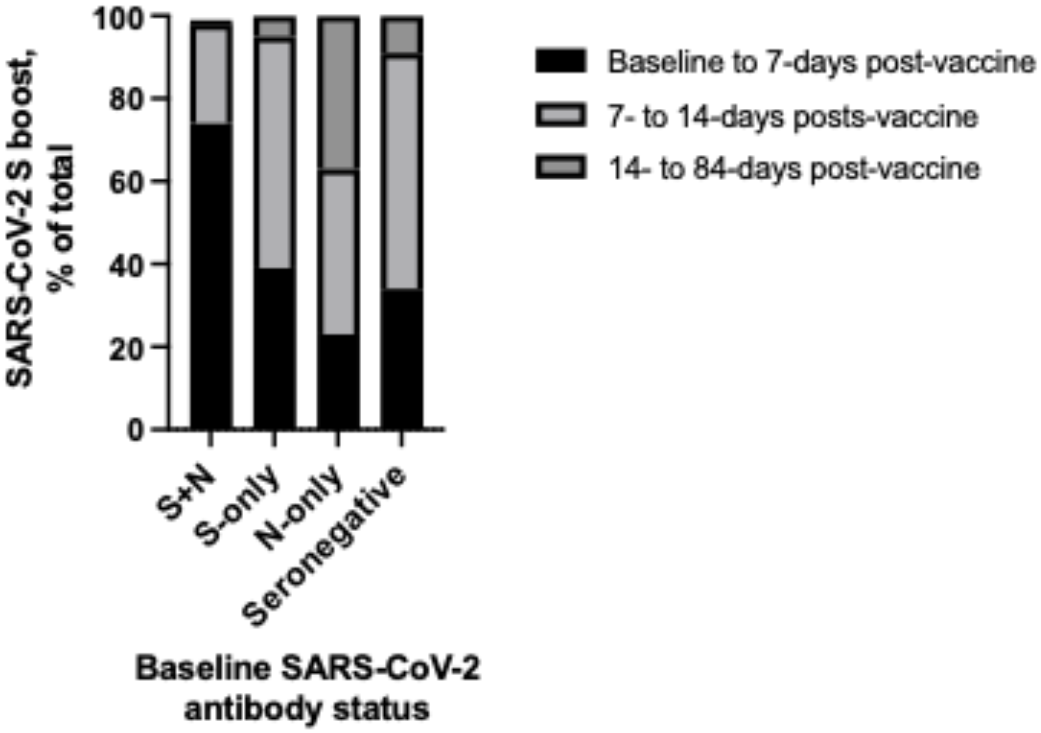
Boosting of SARS-CoV-2 spike (S) in individuals in the general population post-vaccination. Sera sequentially collected from individuals in the general population were subjected to Western blot analysis. The level of antibody boosting was determined by subtracting the mean SARS-CoV-2 S antibody signal between baseline and 7-days post-vaccination, 7- and 14-days post-vaccination, and 14- and 84-days post-vaccination. The S signals from each time period were then summed and plotted as a % of total in a stacked format.

We then examined the T cell response against SARS-CoV-2 N in individuals in the general population 7-days post-vaccination. In individuals with SARS-CoV-2 S+N antibodies, 55.9% (33/59) had a T cell response against SARS-CoV-2 N (Fig. 4A). In individuals with N-only antibodies 100% (2/2) had T cell responses against SARS-CoV-2 N, respectively (Fig. 4B). In individuals who were seronegative, 100% (18/18) did not have T cell responses against SARS-CoV-2 N (Fig. 4C).

**Fig. 4.**
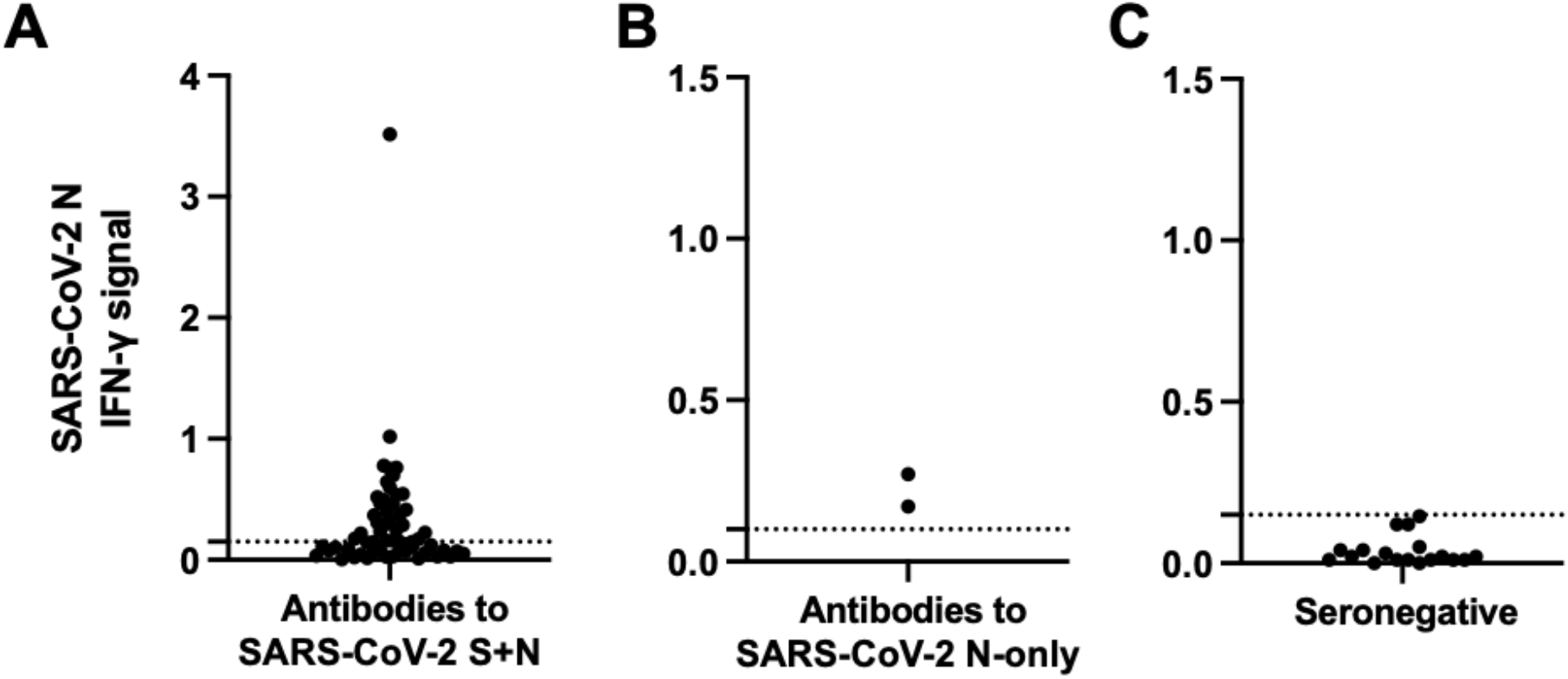
T cell responses to SARS-CoV-2 nucleocapsid (N) 7-days post-vaccination in individuals in the general population. Whole blood samples were stimulated with SARS-CoV-2 N, then supernatants were processed via an IFN-γ enzyme-linked immunosorbent assay for individuals in the general population who had antibodies to A) SARS-CoV-2 S+N, B) N-only, or C) were seronegative. Responses are expressed as an arbitrary unit (IFN-γ signal) based on an OD_405_ measurement. Dashed line, assay cutoff.

Finally, we calculated the sensitivity and specificity to determine the efficacy of using T cell IFN-γ against SARS-CoV-2 N as a diagnostic biomarker for exposure. The positive control group consisted of 16 HCWs with previous RT-PCR confirmation of COVID-19 and the negative control group consisted of 26 HCWs and individuals in the general population who were seronegative. The resulting sensitivity was 87.5% and the specificity was 92.3% (Table 2).

**Table 2.** Performance of T cell IFN-γ against SARS-CoV-2 N in control groups.

| Statistic | Value | 95%<br>Confidence Intervals |
| --- | --- | --- |
| Sensitivity | 87.50% (14/16) | 61.65% to 98.45% |
| Specificity | 92.31% (24/26) | 74.87% to 99.05% |
| Positive Predictive Value | 87.50% | 64.60% to 96.41% |
| Negative Predictive Value | 93.10% | 76.56% to 97.78% |

## Discussion

In Africa, the immunological response to SARS-CoV-2 and non-SARS coronaviruses represents a critical research gap that may shed light on the paradoxical high SARS-CoV-2 infection with low mortality rate as compared to other parts of the world. The goal of the present study was to profile the antibody and T cell responses in natural SARS-CoV-2 infection and COVID-19 vaccination in two Nigerian cohorts. There are four important findings from our study. First, antibodies directed against SARS-CoV-2 S+N, suggestive of previous exposure the virus, were observed in a majority of individuals prior to vaccination. These results agree with other SARS-CoV-2 seroprevalence studies, suggesting a very high infection rate by 2021. Second, having antibodies to SARS CoV-2 N-only is suggestive of pre-existing coronavirus immunity, as represented by HCWs and the general population with N-only antibodies prior to vaccination. Third, SARS-CoV-2 S antibody boosting occurs more rapidly in individuals with prior SARS-CoV-2 infection. Finally, T cell IFN-γ against SARS-CoV-2 N is a robust method to detect exposure to the virus as demonstrated by the high detection rate among the HCWs with prior PCR-confirmed COVID-19.

Studies have shown that the antibody response to natural infection by SARS-CoV-2 is highly variable (16). In some cases, individuals never produce a detectable antibody response, or low antibody titers have been observed in individuals with mild or asymptomatic infections (16, 17). The vast majority of antibody studies have utilized ELISA assays to recombinant SARS CoV-2 antigens. Multiple studies now indicate reduced specificity of these assays when analyzing samples originating from sub-Saharan Africa (11, 18). Of note, our study employed virion based immunoblot which enabled simultaneous visualization of antibody responses to multiple SARS-CoV-2 antigens with multiple positive and negative controls to ensure specificity.

Our antibody data showed a high SARS-CoV-2 seroprevalence in Nigeria with strong antibody reactivity. A majority of individuals, including 71.6% of HCWs and 54.8% of the general population vaccine recipients, had antibodies directed against SARS-CoV-2 S+N (Table 1). These data are in comparison to studies conducted in Nigeria between June and December 2020 showing lower seroprevalence rates ranging between 17-25% (19, 20).

While COVID-19 vaccines have also been shown to induce robust antibody responses, there is strong evidence that these responses decline in the months following the second or boosting doses (21, 22). Our data demonstrate that a majority of general population vaccine recipients, regardless of baseline antibody status, developed SARS-CoV-2 S antibodies post-vaccination (Fig. 2). However, six individuals (5.2% across vaccine population) failed to develop an antibody response even after two vaccine rounds (Fig. 2D-E). Moreover, previous SARS-CoV-2 immunity appears to impact the level of S antibody boosting. Vaccination with the Oxford/AstraZeneca COVID-19 vaccine boosted 74% of S antibodies within 7 days in individuals who already had SARS-CoV-2 S+N antibodies at baseline (Fig. 3). In contrast, only 39%, 23%, or 34% of S boosting occurred within 7 days in individuals who demonstrated S-only or N-only antibodies or were seronegative at baseline, respectively (Fig. 3). Importantly, analysis was not performed on samples beyond 84-days post-vaccination; therefore, we are unable to postulate on antibody waning. However, several studies have demonstrated substantial antibody waning after vaccination with the Oxford/AstraZeneca COVID-19 vaccine (23).

During infection with SARS-CoV-2, S and N proteins are major targets of both antibodies and T cells (24). While SARS-CoV-2 S protein is more genetically diverse among coronavirus infected humans and animals, the N protein is highly conserved (25). Our antibody data suggests pre-existing coronavirus immunity with 10.4% (14/134) of HCWs and 20% (23/115) of individuals in the general population who only had SARS-CoV-2 N antibodies prior to vaccination. These results recapitulate other studies highlighting a similar phenomenon (11, 18, 26). SARS-CoV-2 N seropositivity could suggest undocumented infections with other as yet unrecognized coronaviruses, including animal coronaviruses. There is 93-100% homology between SARS-CoV-2 and several bat and pangolin N proteins at the amino acid level, and both alpha and betacoronaviruses have been isolated from animals in Central and West Africa (11, 27, 28). The dynamics of antibody waning post natural infection is not well studied. It is possible that waning of S antibodies may result in the SARS-CoV-2 N antibody only profile. Prospective serologic studies are needed in these populations to further elucidate this finding.

We also report, for the first time, T cell responses against SARS-CoV-2 N in West Africans. The T cell assay used in this study does not require peripheral blood mononuclear cell (PBMC) isolation, offering ease-of-use and a more time-sensitive procedure from sampling to result. Instead, an individual’s whole blood is collected in lithium heparin tubes, followed by T cell stimulation in the kit’s specialized tubes coated with SARS-CoV-2 N protein or positive/negative controls. After stimulation, supernatants are processed via a one-step ELISA to measure IFN-γ secreted by T cells that responded to the SARS-CoV-2 N protein.

Evidence suggests that most individuals infected with SARS-CoV-2 generate IFN-γ-producing T cells that can be detected in peripheral blood as early as 2-4 days from the onset of symptoms or between 7-14 days after vaccination (29, 30). Our results demonstrate a high sensitivity and specificity in detecting exposure to SARS-CoV-2 based on whole blood IFN-γ. In HCWs with previous RT-PCR confirmation of COVID-19, the sensitivity of T cell IFN-γ against SARS-CoV-2 N was 87.5%. The specificity of the assay in individuals who were seronegative was 92.3% (Table 2). In individuals from the general population who were seronegative, none had detectable T cell IFN-γ responses. However, two HCWs who were seronegative had a positive T cell response. These may have been false positives or the T cell response to SARS-CoV-2 N was skewed by pre-existing immunity to other pathogens including globally endemic human non-SARS coronaviruses (25).

This study has limitations. Our study populations of HCWs and the general population of vaccine recipients were relatively small. Second, while our results are suggestive of pre-existing coronavirus immunity, it was not within the scope of the study to confirm the specific coronavirus responsible for the response. Studies will be required to determine whether pre-existing coronavirus immunity may alter susceptibility to SARS-CoV-2 infection and/or COVID-19 severity. Third, the focus of the T cell response was on SARS-CoV-2 N. While we observed robust responses to N, there are other proteins that have been shown to be targeted by T cells, which may correlate with disease severity. Finally, due to the limited amount of blood collected for each patient, we were unable to define the CD4- and CD8-specific T cell responses.

In conclusion, our study demonstrates high SARS-CoV-2 seroprevalence as well as possible preexisting coronavirus immunity in the Nigerian population in 2021, and also provides new data on T cell responses as well as antibody boosting after vaccination based on prior infection/exposure. Our findings highlight the need for further investigations to better understand the immune mechanisms and consequences related to pre-existing coronavirus immunity in West Africa. Pre-existing B cell and/or T cell memory may have important implications for natural infection and disease outcomes. Identification of conserved antibody and/or T cell epitopes may hold promise for improved vaccines protecting against current and future coronaviruses.

## Data Availability

All data produced in the present study are available upon reasonable request to the corresponding author.

## Acknowledgments

We thank the many healthcare workers at Lagos University Teaching Hospital that participated in this study. We also acknowledge the collaboration with the Lagos State COVID-19 Taskforce in the evaluation of COVID-19 vaccine immunity. This study was funded by Harvard University’s Motsepe Presidential Research Accelerator Fund for Africa.

## Author Contributions

Conceptualization: SA, BBH, PJK. Methodology: SA, BBH, BC, SO, AO, FO, SJO, IEA, AN, DJH, CAC, PJK. Validation: SA, BBH, BC, SO, AO, FO, SJO, IEA, AN, DJH, CAC, PJK. Formal Analysis: SA, BBH, BC, AO, DJH, CAC, PJK. Data Curation: SA, BBH, BC, SO, AO, FO, SJO, IEA, AN, DJH, CAC, PJK. Writing – Original Draft Preparation: BBH. Writing – Review & Editing: SA, BBH, BC, SO, AO, FO, SJO, IEA, AN, DJH, CAC, PJK. Visualization: BBH, BC, CAC, PJK. Supervision: SA, AO, PJK. Funding Acquisition: PJK.

